# SelectStitch: Automated Frame Segmentation and Stitching to Create Composite Images from Otoscope Video Clips

**DOI:** 10.1101/2020.08.12.20173765

**Authors:** Hamidullah Binol, Aaron C. Moberly, M. Khalid Khan Niazi, Garth Essig, Jay Shah, Charles Elmaraghy, Theodoros Teknos, Nazhat Taj-Schaal, Lianbo Yu, Metin N. Gurcan

**Affiliations:** Center for Biomedical Informatics, Wake Forest School of Medicine, Winston-Salem, NC, USA; Department of Otolaryngology, Ohio State University, OH, USA; Case Western Reserve University School of Medicine, OH, USA; University Hospitals Seidman Cancer Center, OH, USA; Department of Internal Medicine, Ohio State University College of Medicine, OH, USA; Department of Biomedical Informatics, Ohio State University, OH, USA

**Keywords:** Computer-assisted Diagnosis, Convolutional Neural Networks, Eardrum abnormalities, Image Stitching, Otoscope, Semantic Segmentation

## Abstract

**Background and Objective:** The aim of this study is to develop and validate an automated image segmentation-based frame selection and stitching framework to create enhanced composite images from otoscope videos. The proposed framework, called SelectStitch, is useful for classifying eardrum abnormalities using a single composite image instead of the entire raw otoscope video dataset.

**Methods:** SelectStitch consists of a convolutional neural network (CNN) based semantic segmentation approach to detect the eardrum in each frame of the otoscope video, and a stitching engine to generate a high-quality composite image from the detected eardrum regions. In this study, we utilize two separate datasets: the first one has 36 otoscope videos that were used to train a semantic segmentation model, and the second one, containing 100 videos, which was used to test the proposed method. Cases from both adult and pediatric patients were used in this study. A configuration of 4-levels depth U-Net architecture was trained to automatically find eardrum regions in each otoscope video frame from the first dataset. After the segmentation, we automatically selected meaningful frames from otoscope videos by using a pre-defined threshold, i.e., it should contain at least an eardrum region of 20% of a frame size. We have generated 100 composite images from the test dataset. Three ear, nose, and throat (ENT) specialists (ENT-I, ENT-II, ENT-III) compared in two rounds the composite images produced by SelectStitch against the composite images that were generated by the base processes, i.e., stitching all the frames from the same video data, in terms of their diagnostic capabilities.

**Results:** In the first round of the study, ENT-I, ENT-II, ENT-III graded improvement for 58, 57, and 71 composite images out of 100, respectively, for SelectStitch over the base composite, reflecting greater diagnostic capabilities. In the repeat assessment, these numbers were 56, 56, and 64, respectively. We observed that only 6%, 3%, and 3% of the cases received a lesser score than the base composite images, respectively, for ENT-I, ENT-II, and ENT-III in Round-1, and 4%, 0%, and 2% of the cases in Round-2.

**Conclusions:** Frame selection improves the diagnostic quality of composite images from otoscope video clips.

## 1. Introduction

Ear infections, particularly acute infections of the middle ear (i.e., acute otitis media – AOM), are a major health problem in the pediatric population [1]. Otoscope is used in the clinical examination of the eardrum or tympanic membrane (TM, an organ that separates the ear canal from the middle ear) as the basic diagnostic apparatus for checking the status of the ear canal and TM. Nevertheless, both clinician and computerized system diagnostic accuracies are heavily influenced by the limitations of otoscopy, such as small field of view [2], poor illumination, or partial occlusions [3], e.g., by hair or wax. Clinicians have around 75 percent of diagnostic accuracy [4-8] with viewing single otoscopic images grabbed from digital otoscopes.

Most of the computer-assisted methods in this field analyze two-dimensional images captured by traditional otoscopes and oto-endoscopes. Unfortunately, these methods can only distinguish among a limited number of TM abnormalities. For instance, Kuruvilla et al. proposed a method to distinguish AOM from other abnormalities like otitis media with effusion (OME) [5, 9]. However, it is difficult to generalize their approach to more than two categories because they need to design more handcrafted features to identify other TM abnormalities such as TM perforation (a hole in the eardrum), TM retraction (a condition in which a part of the eardrum lies deeper within the ear than its normal position), and tympanosclerosis (a condition including scarring or accumulation of calcium deposits within the TM).

To explore the computer-assisted detectability of a wide range of eardrum abnormalities, we employed both deep learning techniques [10] and traditional approaches that require hand-crafted features [11, 12] in our previous works. While prior studies reported promising results, they rely on a single image rather than raw video for eardrum abnormalities [13]. Manually selecting a representative frame from even a few seconds of those videos is extremely time consuming and subject to high inter- and intra-reader variability. The complex topology of the eardrum requires multiple images at varying depths of focus to properly capture the eardrum. For this reason, even if a best frame could be selected from the video, it might not contain the entire view of the TM. Therefore, a clear and comprehensive composite image generated automatically from otoscope video clips would be helpful for accurate and automated diagnosis of eardrum abnormalities.

The aim of this study is to develop and validate an automated image segmentation and stitching framework, called *SelectStitch*, to create enhanced composite images from otoscope videos. For this purpose, a semantic segmentation-based framework is proposed. The segmentation and subsequent stitching enable us to automatically select meaningful frames from otoscope videos and reduce irrelevant ones (e.g., those heavily blurred with excessive amount of cerumen). A meaningful frame is a frame that contains a specific proportion of an eardrum. We propose a modified U-Net [14] based semantic segmentation approach to identify meaningful frames, which are used by our stitching engine to create a composite image. We then compare the diagnostic decisions of three ear, nose, and throat (ENT) physicians after reviewing these new composite images, relative to the images generated by using the entire frames of the video recorded by traditional handheld otoscopy (i.e. composite images generated without frame selection). Fig. 1 gives an overview of our complete analysis pipeline in this study.

**Fig. 1.**
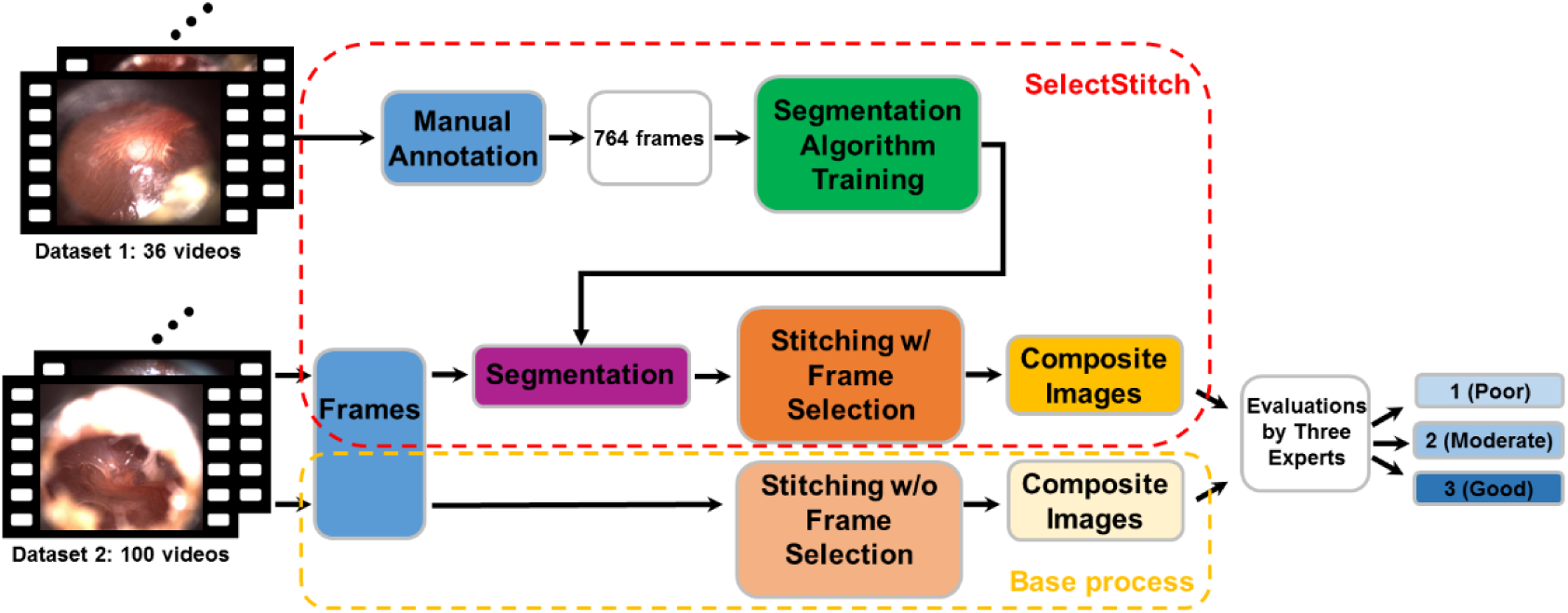
Overview of our study structure. 36 otoscope videos were used for training a segmentation algorithm which is then used to select appropriate frames for the 100 otoscope test videos. The composite videos from a control set without frame selection and SelectStitch method are then compared visually by three ENTs in two rounds of evaluation.

The rest of this paper is organized as follows: Section 2 describes the data and the proposed framework. Experimental results are presented in Section 3. The discussion and concluding remarks are respectively included in Sections 4 and 5.

## 2. The proposed methodology

### 2.1 Materials

A centralized database of high-resolution digital adult and pediatric images was created for this particular project, captured at Ear, Nose, and Throat (ENT) clinics and primary care settings at the Ohio State University (OSU) and Nationwide Children’s Hospital (NCH) in Columbus, Ohio, USA in accordance with an Institutional Review Board (IRB) approved protocol. A high definition (HD) video otoscope (JEDMED Horus+ HD Video Otoscope, St. Louis, MO) was utilized to capture and record the video data. The video frames are of size 1440 by 1080 pixels and are recorded in MOV format.

Our dataset included 136 otoscope videos. We divided these into two separate datasets: the first one (Dataset 1) with 36 otoscope videos were used to develop the semantic segmentation model, and the second one (Dataset 2) containing 100 videos for an independent set was used for employing the stitching process along with a reader study to assess the efficiency of the proposed method. In Dataset 1, the eardrum regions in 764 frames were manually annotated by two otolaryngologists (ACM and CE) (Fig. 2). Table 1 shows the distribution of major diagnostic categories as reported by the expert physicians.

**Fig. 2.**
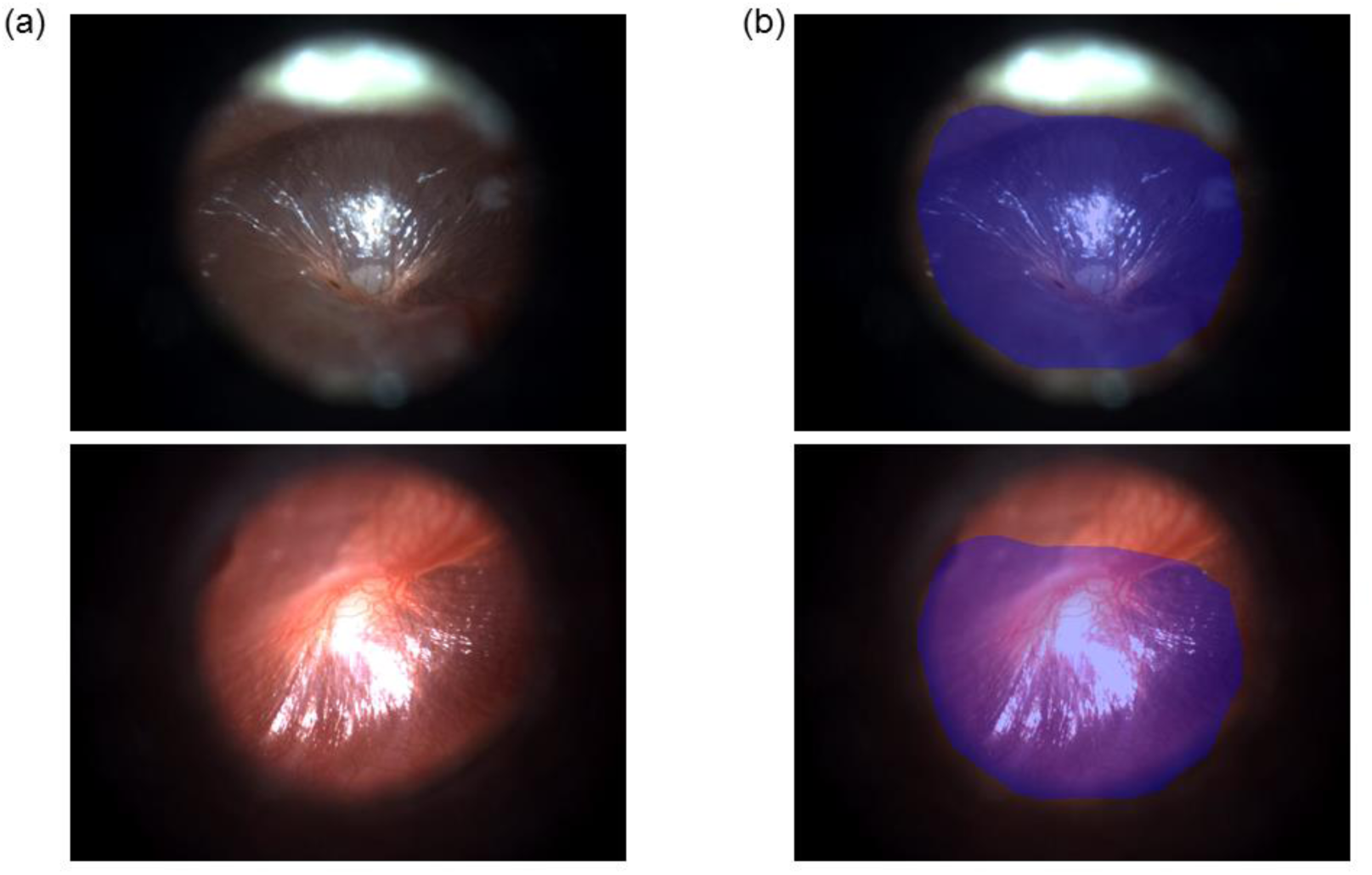
Examples of annotation. (a) Original tympanic otoscopy images; (b) annotated images. Glare and blur are excluded from the target area by the annotator.

**Table 1.** Number of Abnormalities in Dataset 2.

| **Abnormalities** | **Number of Occurrences** |
| --- | --- |
| Effusion | 27 |
| Tympanosclerosis | 22 |
| Perforation | 15 |
| Retraction | 12 |
| Acute otitis media (AOM) | 11 |
| Cholesteatoma | 7 |
| Other minor cases | 6 |
| **Total** | **100** |

### 2.2 U-Net based semantic segmentation

U-Net was introduced in biomedical imaging to improve precision and localization of microscopic images of neuronal structures. The architecture builds upon the fully convolutional network [15] and is similar to the deconvolutional network [16]. In a deconvolutional network, a stack of convolutional layers — where each layer halves the size of the image but doubles the number of channels — encodes the image into a small and deep representation. That encoding is then decoded to the original size of the image by a stack of up-sampling layers. The U-Net adds additional skip connections between layers at the same hierarchical level in the encoder and decoder. This allows low-level information to flow directly from the high-resolution input to the high-resolution output.

We trained a U-Net architecture with an encoder depth of four to automatically find eardrum regions in each otoscope video frame. In order to reduce the effects of overfitting, we took advantage of data augmentation [17], which involves random horizontal flips, image sharpener, affine transformations between −45 and 45 degrees, and elastic transformations with three different *α* intervals of (45, 50), (55, 60), and (65, 80) with σ = 5 as described in [18]. The augmentation process was implemented using the *imgaug* library [19]. Then, to scale inside the image randomly, images were shrunk and enlarged within a range of (−0.5, +0.5). After data augmentation, we had 15,280 additional images, i.e., generated 20 augmented images for each of the 764 images in Dataset 1, to train the segmentation network.

The performance of the segmentation algorithm was computed in a k-fold cross-validation [20] (with *k* = 10). It should be emphasized that the segmentation framework was exposed neither to the validation images (Dataset 2) nor the augmented images resulting from the validation set.

### 2.3 Image stitching

Image stitching was performed with an OpenCV image-processing library for a stepwise process of correcting optical distortion, cropping the circular TM field of view, and estimating affine transforms between neighboring fields using Speeded Up Robust Features (SURF) [21] key points matching. These transforms describe the translation, rotation, and skew of each peripheral field relative to the central field. Finally, a full-resolution mosaic is generated using the estimated transforms, and overlapping regions are linearly blended.

To save time on developing a customized image stitching algorithm and to focus on the clinical impact of the proposed framework, we used Image Composite Editor (ICE) 2.0 [22] software package, created by the Microsoft Research Computational Photography Group, which generates seamlessly combined composite images. Microsoft ICE is distributed as a freeware for non-commercial use.

Microsoft ICE does not have the capability of stitching images from totally different scenes. Therefore, reducing TM-irrelevant frames, as proposed in this study, was needed to utilize this tool. Our solution provides a feasible and practical alternative to manual selection. The proposed framework, SelectStitch, consists of two main processes, i.e., semantic segmentation and image stitching, as depicted in Fig. 3.

**Fig. 3.**
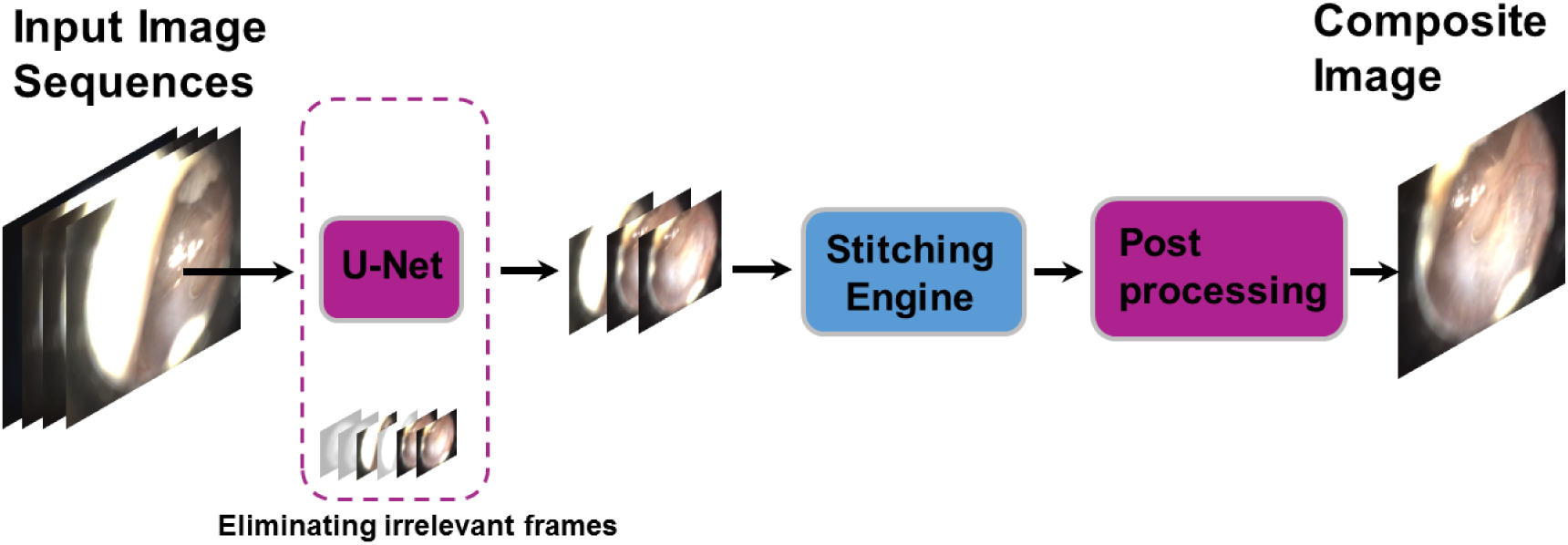
Proposed framework for composite images generation from otoscope video clips.

### 2.4 Post-processing

Post-processing consisting of two steps, cropping and image enhancement, was applied to the output of the composite image generator. Most composite images produced by the ICE included black background areas, which were redundant and removed after blurring with a Gaussian filter [23], gray level thresholding [24] and foreground detection on the thresholded, binary image.

An image captured in an outdoor scene could be highly degraded due to poor-or over lighting conditions or if there are different suspension particles like water droplets or dust particles. These particles may cause the irradiance coming from the object to be scattered or absorbed, leading to haze, smoke or fog. The resulting images are degraded, and the color and contrast are shifted from the original irradiance at the time of capture of the image. The image needs to be de-hazed before it can be analyzed. While our ear images were not captured outdoors, the inverted low-light images common in eardrum imaging results in hazy images. Therefore, we borrowed a de-hazing technique [25] to solve low-light image condition in this study. Fig. 4 gives the before and after images for a sample output of the cropping and dehazing technique.

**Fig. 4.**
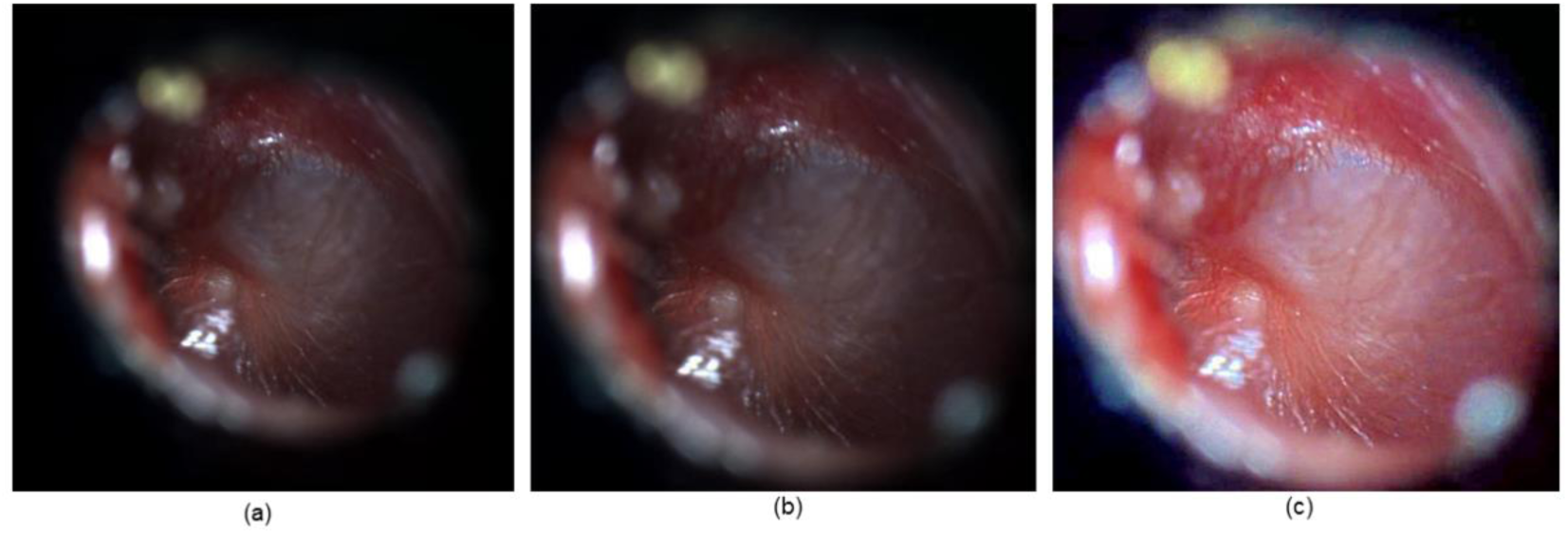
Example result of post-processing steps (a) Original image generated by the composite image generator; (b) Image cropped by applying converting to gray level, blurring with a Gaussian filter, gray level thresholding and identification of the coordinates of the area that is needed for subsequent processing; (c) Image enhanced by a de-hazing technique.

### 2.5 Experimental setting

Segmentation experiments were conducted on Wake Forest Baptist Medical Center’s high-performance computer cluster. We took advantage of 16GB NVIDIA Tesla P100 PCI-E GPU. We used the Deep Learning Toolbox of MATLAB R2018b to implement the U-Net architecture, and Dice coefficient [26, 27] to evaluate the performance of the segmentation.

Minimization of the loss is achieved via stochastic gradient descent using the Adam optimizer [28] and learning rate of 0.001. We used mini-batches of size 16 for U-Net. Early stopping was employed to avoid over-fitting [29, 30].

In this study, we generated 100 composite images from the corresponding videos in Dataset 2. The videos in Dataset 2 were categorized into different eardrum abnormalities by four expert physicians (three ENT specialists and one pediatrician) based on physical examination and evaluation of the patient at the time of the encounter.

#### Base process

In the base-process, we stitch all the frames of the video data regardless of their content to create composite images. The base process does not incorporate neither a human intervention nor a computerized technique to process or analyze video frames before stitching step, i.e. every frame is used in the resulting composite image. Unlike SelectStitch, the resulting composite images do not benefit from segmentation-based frame selection nor post-processing.

#### Two rounds of evaluation

The composite images produced by SelectStitch were compared by three ENT specialists against the base processes in terms of their diagnostic capabilities. This reader study provides an evaluation of a new measurement or analysis method [31]. In order to do this, three ENT specialists graded each composite image using one of the three categories: 1 (Poor) refers to a situation that he/she cannot diagnose well or the image has serious problems; 2 (Moderate) refers to an image which is poor quality but still usable for diagnosis; and finally 3 (Good) refers to a good quality composite image which can be used for diagnosis. Doing these measurements with three ENTs provided us a measure of inter-reader variability, and repeating the study in a second round provided us an estimate of intra-reader variability.

### 2.6 Statistical analysis

To evaluate changes in scoring the composite images generated by SelectStitch compared to the base process images in two rounds of assessments of three ENT specialists, we employed an ordinal logistic regression model on image scoring (possible levels of 1, 2, or 3) to test the differences between two methods (i.e. base process and SelectStitch) and two rounds. To evaluate the inter-reader variability within rounds and the intra-reader variability between rounds for each method, we calculated Kendall’s coefficient of concordance, which ranges between 0 (representing no agreement) and 1 (representing perfect agreement).

## 3. Results

We trained U-Net to segment each frames of otoscope videos. Using 10-fold cross validation, we noted Dice coefficient of 0.84 ± 0.03. While investigating the effect of the segmentation process (as recommended by Zijdenbos et al. [32]) was beyond the scope of this study, dice coefficient of greater than 0.70 generally indicates a good overlap. At test time for composite image generation, a frame is considered relevant if at least it contains an eardrum region of an experimentally determined threshold of 20%, of its size. TM-relevant frames were utilized to generate composite images (see the SelectStitch process in Fig. 1).

Table 2 gives the changes in scoring the composite images generated by SelectStitch compared to the base images among two rounds of assessments. An examination of Table 2 shows that when SelectStitch is used, the readers gave fewer responses of a score of 1 (Poor) and more responses of a score of 3 (Good) compared to the base composite images, regardless of round.

**Table 2.** Changes in Scores on the Evaluation of SelectStitch Composite Images Compared to Base Process Composite Images over Two Rounds for Three Experts where the Symbol of > Means “to”; the Symbol of ↔ Represents No Change Cases.

| Reader | Round-1 |  |  |  |  |  |  | Round-2 |  |  |  |  |  |  |
| --- | --- | --- | --- | --- | --- | --- | --- | --- | --- | --- | --- | --- | --- | --- |
| | Improvements | | | Deterioration | | | ↔ | Improvements | | | Deterioration | | | ↔ |
|  | 1 > 3 | 1 > 2 | 2 > 3 | 3 > 1 | 2 > 1 | 3 > 2 |  | 1 > 3 | 1 > 2 | 2 > 3 | 3 > 1 | 2 > 1 | 3 > 2 |  |
| ENT-I | 8 | 32 | 18 | 0 | 0 | 6 | 36 | 13 | 22 | 21 | 0 | 0 | 4 | 40 |
| ENT-II | 24 | 15 | 18 | 0 | 1 | 2 | 40 | 17 | 29 | 10 | 0 | 0 | 0 | 44 |
| ENT-III | 17 | 24 | 30 | 0 | 1 | 2 | 26 | 13 | 40 | 11 | 0 | 0 | 2 | 34 |

Table 2 indicates that using SelectStitch composite images, ENT-I improved 32 images from category 1 (Poor) to 2 (Moderate), 18 images from 2 to 3 (Good), and 8 composite images from category 1 to 3; ENT-II improved 15 composite images from 1 to 2, 18 images from 2 to 3, and 24 composite images from category 1 to 3 and ENT-III improved 24 composite images from 1 to 2, 30 images from 2 to 3, and 17 composite images from category 1 to 3 for Round-1.

To illustrate the effectiveness of our proposed framework for otoscope video stitching, we presented the composite images of three otoscope video clips in Fig. 5.

**Fig. 5.**
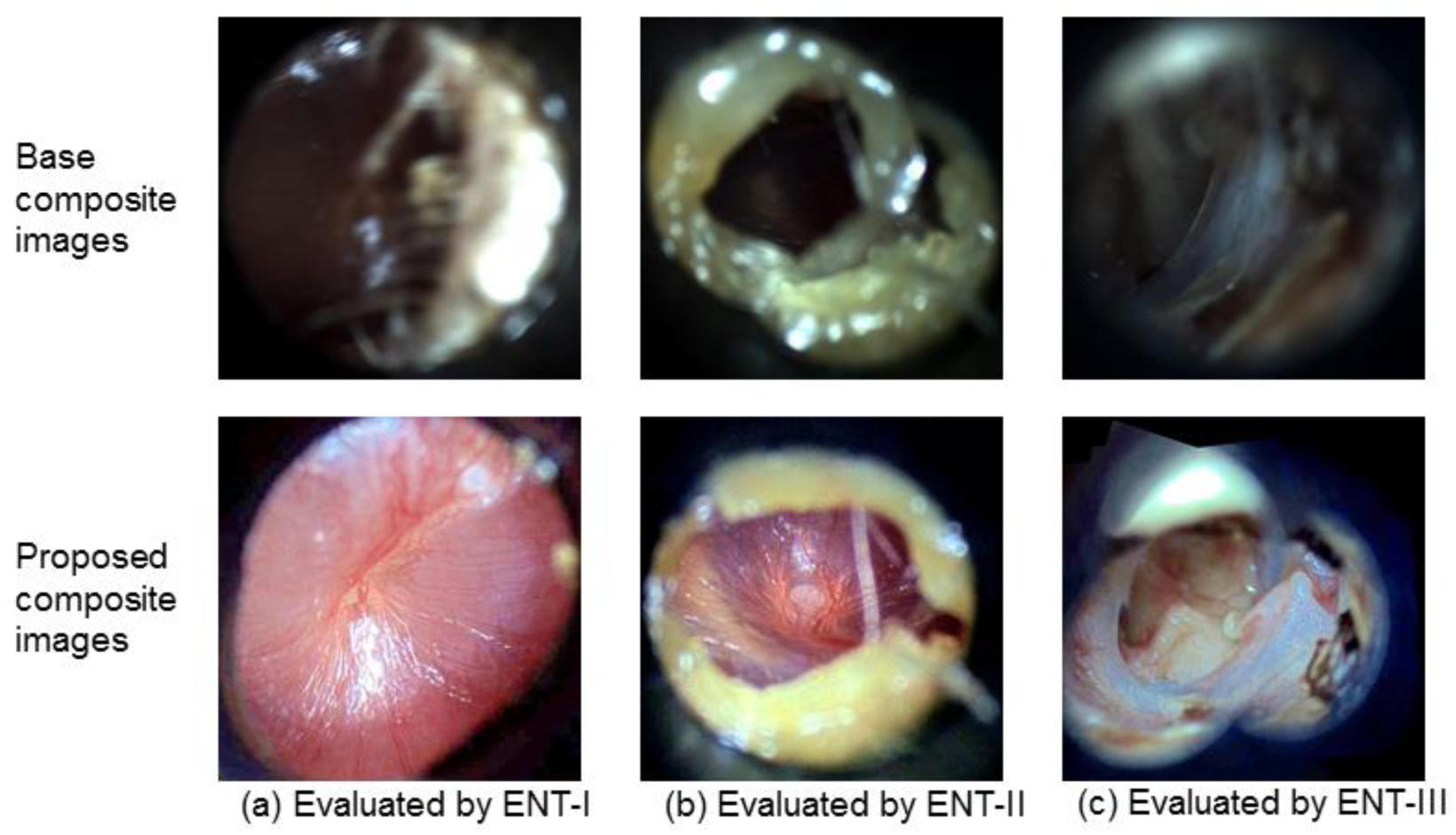
Examples of successful composite results using our approach. The first row shows the images generated by the base processes and the second row shows the images by our approach, SelectStitch. Each column indicates the number of readers. Each reader gives the score of 1 for the base image and 3 for the proposed image.

We used an ordinal logistic regression model to compare image scoring between SelectStitch and the base method and between two rounds. There is a significant difference in scoring between the base and proposed methods at p value of 0.0007, and the base method has 6.5 times higher risk of scoring less than the proposed method.

The probabilities on scoring are listed in Table 3. At Round-1, the base method has probability of 0.43 for scoring below 2 and has probability of 0.84 for scoring below 3, but the proposed method has probability of 0.10 for scoring below 2 and has probability of 0.44 for scoring below 3. At Round-2, the base method has probability of 0.52 for scoring below 2 and has probability of 0.88 for scoring below 3, but the proposed method has probability of 0.14 for scoring below 2 and has probability of 0.53 for scoring below 3. In summary, the base method has higher probability for scoring less than the proposed method at both rounds.

**Table 3.**
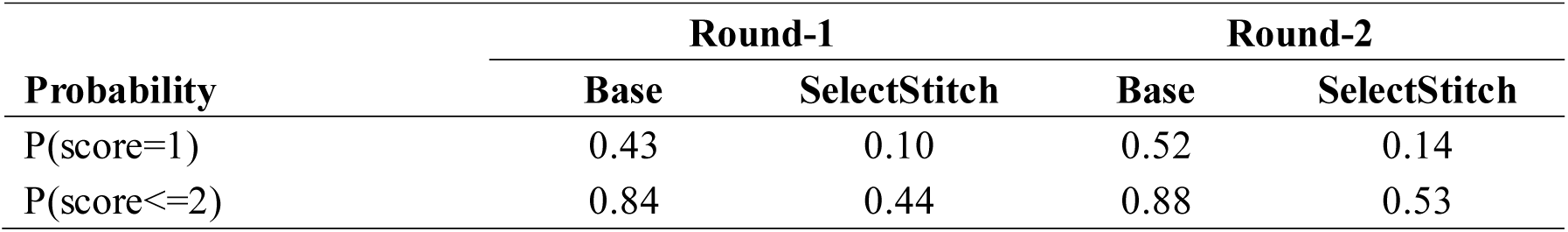
Probability on Scoring.

As with other medical tests, there are intra- and inter-reader variations in assessing the eardrum abnormalities. Table 4 gives the inter- and intra-reader variability among the rounds for each method (values < 0 as indicating no agreement and 0 – 0.20 as slight, 0.21 – 0.40 as fair, 0.41 – 0.60 as moderate, 0.61 – 0.80 as substantial, and 0.81 – 1 as almost perfect agreement). We observed the base method has higher inter- and intra-reader agreement in assessment of evaluation of composite images according to their diagnostic capabilities. This could be explained by the fact that the ENTs agreed more on the poor quality of cases than deciding whether the improvement category belongs to 2 or 3.

**Table 4.**
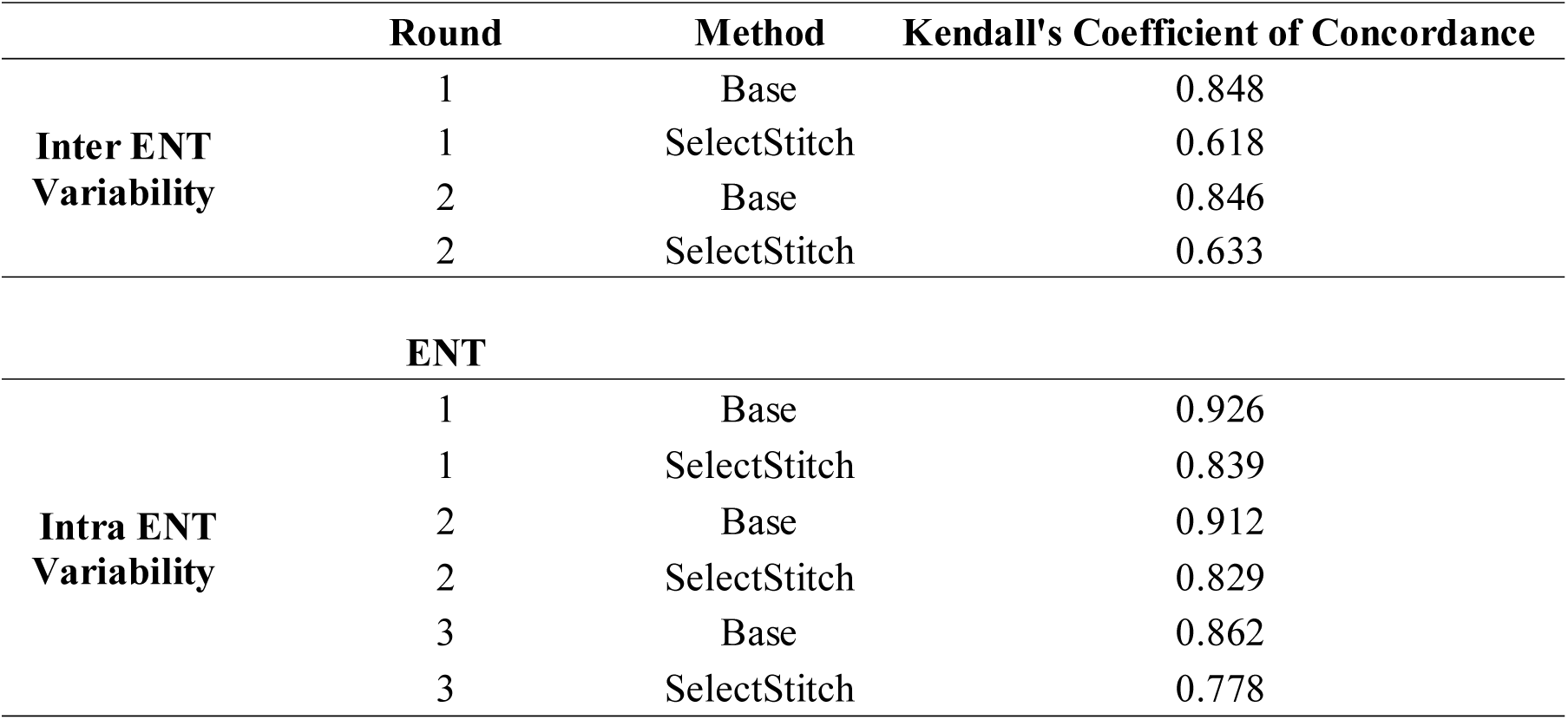
Inter- and Intra-ENT Variability.

## 4. Discussion

To the best of our knowledge, this is a first attempt at representing an otoscope video with a single composite image while preserving its diagnostic capability as much as possible. Three ENT experts evaluated the images in the scale of poor, moderate, and good and the results (see Table 2) show that, on average among experts and evaluation rounds, in 60.3% of the cases the diagnostic quality improved and in 15.3% of the cases this improvement was from poor to good and 18.0% from moderate to good. Only 3.0% of the cases deteriorated: 2.7% from good to moderate and only 0.3% from moderate to poor. There is a statistically significant difference in scoring between the base and proposed methods at p value of 0.0007.

Exemplary composite images for the case of same scoring (score of 2) are shown in Fig 6. In Fig. 6, each column illustrates the evaluations from different readers. As can be observed in Fig. 6, it is likely that better scores for the proposed images will be obtained in a wider assessment scale (e.g. 1-5).

**Fig. 6.**
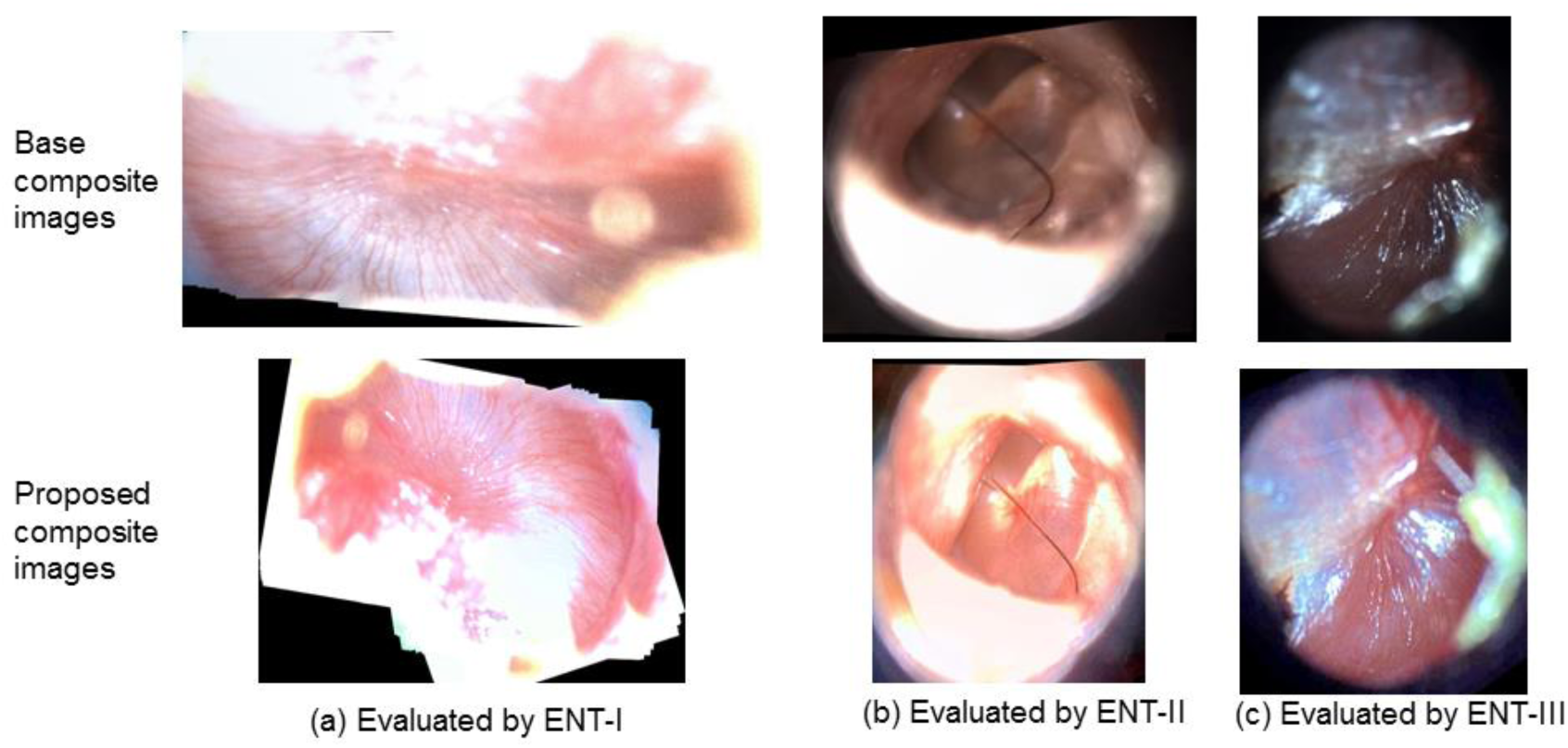
Examples of the same evaluation score for both composite images. The first row shows the images generated by the base processes and the second row shows the images by SelectStitch. Each column indicates also the number of reader and each reader gives the score of 2 for both base and the proposed image.

Exemplary composite images for the cases where the results deteriorated after SelectStitch are shown in Fig. 7, in which the proposed images were consistently scored as poorer than the base images. The comment noted by the ENT-I for the first column was that the base image looks more natural and the proposed image has an unnatural lighting, making difficult to see the disease, which is retraction. The drawback for the composite image generated by the proposed framework (see the second column of Fig. 7) was overexposing, but the base image received the score of 3 although it is also mentioned that it has partial view. For the third column, ENT-III noted that the SelectStitch image has glare. We also noticed a particular SelectStitch composite image (see fourth and fifth column in Fig. 7) that was given a score of 2, respectively, by ENT-I and ENT-III in both rounds while the base score was 3. For the fourth column in Fig. 7, the sample was having multiple abnormalities, i.e., Perforation, Prosthesis, and Tympanosclerosis; and the reader noted that proposed composite image has “a little unnatural lighting.” For the fifth column, the sample was originally labeled as Perforation, Cholesteatoma, and Tympanosclerosis; and the reader evaluated the proposed image as “a little blurry.” For these cases, we can conjecture that the calcification and scarring present in Tympanosclerosis made the generation of enhanced composite image with natural looking colors even more difficult.

**Fig. 7.**
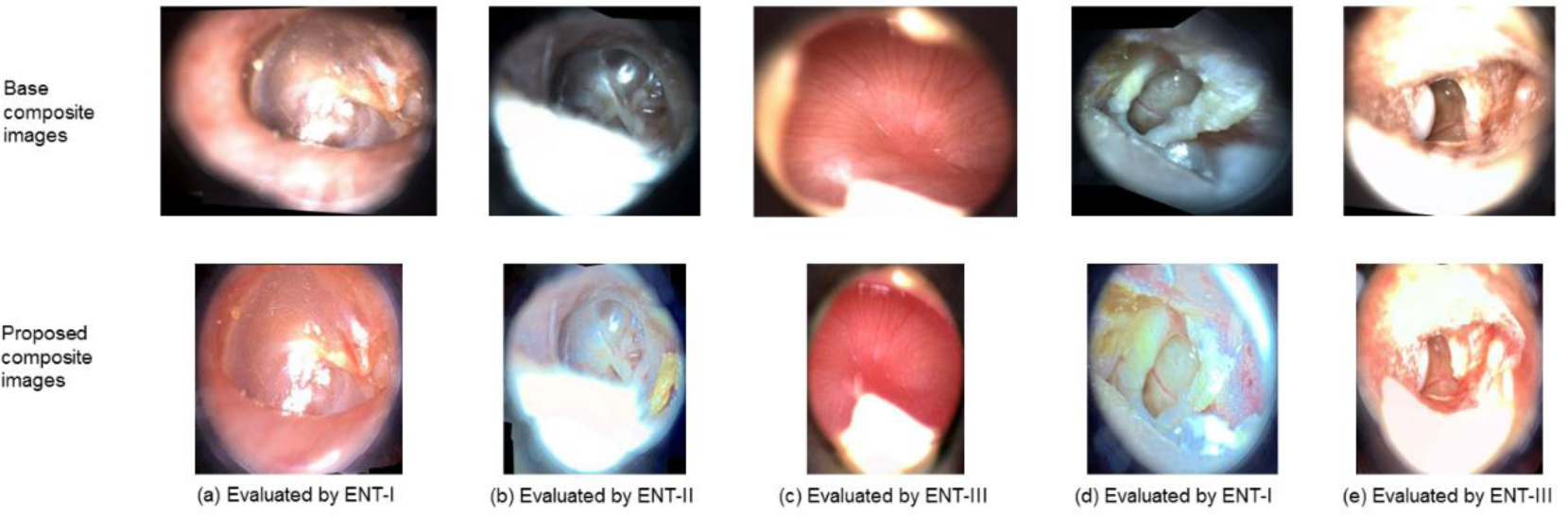
Exemplary composite images for the cases where the results deteriorated after SelectStitch. The first row shows the images generated by the base processes and the second row shows the images by SelectStitch. In all cases, the readers gave a score of 3 for base process composite image and 2 for the SelectStitch composite image.

We also observed that there are six, three, and three cases which ENTs give less scores compared to the base composite images in Round-1 and four, zero, and two cases in Round-2. We noticed that the composite images resulting from the proposed framework that received a lesser score than the base composite images suffered from over-exposure as the dehazing algorithm failed to preserve the color information. To improve the diagnostic capability of the composite image, it should be enhanced by preserving its original color as much as possible. We also noted that in no evaluation, SelectStitch image received the score of 1 while the base process image score was 3. There was a significant difference in scoring between the first and second rounds at p value of 0.001, and the second round has 1.44 times higher risk of scoring less than the first round. It is possible that ENTs with Round-1 experience might have better judgement at second round.

Although this work provides a proof-of-concept approach to obtain one representative composite image of the TM, the intra-class variability is a substantial challenge. Thus, to obtain a robust segmentation, more training samples from scarred eardrums, e.g., tympanosclerosis are required. With a larger database, a multi-modal image segmentation could also be developed.

There are some limitations to this study. First, the relevant frames were determined according to the amount of eardrum they capture. If the amount of eardrum in a frame is above a certain threshold, then it was considered a relevant frame. While our study shows promising results, we expect that additional criteria could be developed to select the frames (or even parts of the frames) to be stitched. This would increase the probability that all the relevant frames (and parts of the frames) could be used to properly represent pathologies present in eardrums.

In our study, we did not evaluate the diagnostic accuracy of composite images over either single images selected from the video or the entire of otoscopic video. Although, we expect that the proposed composite images will be superior (or at least equal) to the manually selected single images in terms of diagnostic capability, a comparison study is needed but is beyond the scope of the current study and will be the subject of our future studies.

## 5. Conclusions

In this study, we developed a framework to obtain enhanced composite images from otoscope video clips and evaluated its effectiveness with a reader study. The data used in our study were acquired from a hand-held HD video imaging system. The results of this study have shown that an appropriate frame selection applied on otoscopy videos can significantly improve the diagnostic quality of composite images generated by these selected frames. We envision that the proposed approach has the potential to provide valuable diagnostic information in the form of normative TM information.

Our system could be very valuable to provide a comprehensive view of the eardrum to clinicians, supporting more appropriate treatment. These composite images may increase the accuracy and efficiency of automated image analysis systems. Such a system can analyze the composite image, on a cloud-based platform, produced from a video-otoscope taken by a trained healthcare individual. These systems could be particularly helpful in case of shortage of specialists who can accurately diagnose eardrum abnormalities, especially in low- and middle-income countries and/or in rural areas [9]. Moreover, this single image could be stored and transferred between health institutions or computation environments instead of a whole video sequence.

More data is needed to explore the generalization of the proposed framework to different kinds of eardrum abnormalities. For example, some abnormalities (TM with retraction, perforation with discharge and cholesteatoma) are more difficult to diagnose than some particular ones such as clear perforation [33]. A further research study will be needed to build models that could analyze the whole spectrum of otoscopic videos faced in clinical settings.

## Data Availability

The data that support the findings of this study are available from the corresponding author, HB, upon reasonable request.

## Acknowledgments

The project described was supported in part by Award R21 DC016972 (PIs: Gurcan, Moberly) from National Institute on Deafness and Other Communication Disorders. The content is solely the responsibility of the authors and does not necessarily represent the official views of the National Institute on Deafness and Other Communication Disorders or the National Institutes of Health.

## Author Contributions

Conceptualization, Hamidullah Binol, M. Khalid Khan Niazi and Metin N. Gurcan; Data curation, Aaron C. Moberly, Garth Essig, Jay Shah, Charles Elmaraghy, Theodoros Teknos and Nazhat Taj-Schaal; Formal analysis, Lianbo Yu; Funding acquisition, Aaron C. Moberly and Metin N. Gurcan; Investigation, Aaron C. Moberly and Metin N. Gurcan; Methodology, Hamidullah Binol, M. Khalid Khan Niazi and Metin N. Gurcan; Project administration, Aaron C. Moberly and Metin N. Gurcan; Software, Hamidullah Binol; Supervision, Aaron C. Moberly and Metin N. Gurcan; Writing – original draft, Hamidullah Binol; Writing – review & editing, Hamidullah Binol, Aaron C. Moberly, M. Khalid Khan Niazi, Garth Essig, Jay Shah, Charles Elmaraghy, Theodoros Teknos, Nazhat Taj-Schaal, Lianbo Yu and Metin N. Gurcan.

